# Does Accreditation Symbolize Quality in Public Healthcare Delivery? An Investigation of Hospitals in Kerala

**DOI:** 10.1101/2020.08.22.20170837

**Authors:** Sindhu Joseph

## Abstract

Accreditation has become an important benchmark for healthcare organisations, and accordingly, many government hospitals in Kerala got accredited with national level (NABH) and state level (KASH) accreditation programmes. This study examined the quality of public healthcare delivery in these accredited hospitals while having a comparison with the non-accredited hospitals. It also compared the impact of national and state-level accreditation programmes in Kerala public healthcare settings. This cross-sectional study conducted between July 2017 and July 2018, employing a positivist approach using stratified random sampling. In total, 621 samples were collected from in-patients of both accredited (NABH and KASH) (312) and nonaccredited (309) public healthcare institutions in Kerala. Nine constructs overarching the quality of healthcare delivery and patient satisfaction construct are used in the study. The study found that patient satisfaction is identical in both accredited and nonaccredited hospitals (M=4.28). Patient satisfaction in NABH accredited hospital (M=4.27±0.67874) is lower than that of KASH accredited hospital (M=4.30±1.25417). The mean score of six constructs of quality healthcare delivery of KASH accredited hospitals is higher than NABH accredited. Thus, the study concluded that accreditation, regardless of its type, has no impact on patient satisfaction even though the accreditation process slightly improved different dimensions of quality healthcare delivery.

## 1. INTRODUCTION

Internationally, since the 1970s, healthcare accreditation programs and accrediting organisations emerged and developed to enhance the healthcare quality improvement activities (Almoajel, 2012; World Health Organization, 2003). This process includes self-assessment and external peer assessment to assess their level of performance against established standards, protocols, laws and regulations. Accreditation demands commitment from the healthcare organisations to improve quality, patient safety, efficiency and accountability and, therefore, increases public acknowledgement (Pomey et al., 2005;Yousefinezhadi et al., 2020). This process measures quality of healthcare institution using a standardised tool which may include the details of qualification, experience and training of healthcare professionals, patient facilities, patient-staff ratios, and acceptance of medical insurance schemes.

There are programmes at international, national and state levels for accrediting hospitals like Joint Commission International (JCI), National Accreditation Board for Hospitals & Healthcare Providers (NABH) in India and Kerala Accreditation Standards for Hospitals (KASH) in Kerala. NABH is founded by the Government of India in 2006 as a benchmark for excellence in healthcare to establish and operate accreditation programme for healthcare organisations. NABH is a constituent board of Quality Council of India (QCI), set up to establish and operate accreditation programme for healthcare organisations. The programme focuses on continuous quality enhancement in terms of patient safety and healthcare delivery based upon national/international standards and mandates to fulfil and operationalise a comprehensive list of 500+ distinct features. Kerala is the first among the states of India to undergo NABH accreditation of its hospitals.

### 1.1 Accreditation and healthcare of Kerala

According to World Bank and Niti Aayog (India), the health indicators of Kerala are on par with the western countries, and the Kerala healthcare model is accepted globally(Kerala State Planning Board, 2017) especially after successfully managing the epidemic Nipah Fever and the pandemic COVID-19. There are 1,280 health institutions with 38,004 beds and 5,465 doctors under Health Services Department (DHS) consisting 848 Primary Health Centers(PHCs), 232 Community Health Centers(CHCs), 81 Taluk Head Quarters hospitals(THQH), 18 District Hospitals, 18 General hospitals(GHs) and eight Women & Children(W&C) hospitals (National Rural Health Mission, 2013;Maya, 2015). According to the minutes of the 11^th^ Executive Committee meeting of Kerala State Health and Family Welfare Society held on 14^th^ July 2010, ‘quality is a guiding principle’ in assessing how well the health system is performing (National Rural Health Mission, 2013).

Kerala has the largest number of NABH accredited hospitals in India under the public sector (Maya, 2015). The new accreditation program, KASH, was introduced in the year 2012, as an initial level accreditation to enhance the quality of curative and preventive healthcare services with the state-of-the-art technology and implemented in selected PHCs, CHCs, THQHs and higher hospitals (National Rural Health Mission, 2013). In order to uplift the quality standards and services given by the public sector hospitals in all care settings, the criteria for KASH was developed in such a way that implementing the program is probable with a modest investment. After achieving KASH, the hospitals may choose for higher accreditation standards like NABH, which require more investment and effort (National Rural Health Mission, 2013).

### 1.2 Quality and patient satisfaction in healthcare delivery

Many researchers have made seminal contributions on service quality and resulting patient satisfaction. Accreditation Canada defines quality improvement (QI) as “the degree of excellence; the extent to which an organisation meets its clients’ needs and exceeds their expectations” (Mondoux, Calder-Sprackman and Thull-Freedman, 2020, p.11). Previous studies have emphasised varied aspects to study the quality healthcare such as availability, accessibility, affordability, acceptability, appropriateness, competency, effective service delivery, privacy, state-of-the-art technology, care, physical environment, responsiveness, admission, treatment, patient-centeredness, waiting-time, cleanliness and hygiene, attitude of doctors and nurses, reliability, comprehensiveness, continuity and equity (Al Tehewy et al., 2009; Amin and Nasharuddin, 2013; Cheng, 2003; Joseph, 2012, 2016, 2017; Linder-Pelz, 1982; Mosadeghrad, 2014; Peprah, 2014; Saeed and Mohamed, 2002; Tashkandi, Hejazi, and Lingawi, 2017; Ware et al., 1983; Zineldin, 2006).

The outcome of quality healthcare delivery overarching the above dimensions is patient satisfaction. Studies on the above mentioned constructs are there in the international and Indian contexts (Banyai, 2012; Delgoshaei, Ravaghi, and Abolhassani, 2012; Grewal et al., 2012; Kavitha, 2012; Lin, 2004; Newcomer, 1997; Parasuraman et al., 1985; Powell, 2001; Saxena, 2009; Solayappan et al., 2011; Verlinde et al., 2012; Yeoh et al., 2013). Patient satisfaction is an essential factor for maintaining long-term relationships, reflected in revisits and willingness to recommend(Elleuch, 2008; Verlinde et al., 2012). Linder-Pelz (1982, p.14) defined patient satisfaction as an “individual’s positive evaluation of distinct dimensions of healthcare.” A well-designed patient satisfaction survey will combine these elements as it relates to the total patient experience(Powell, 2001).

### 1.3 Impact of accreditation programmes

There exists a considerable body of literature, establishing the positive impact of accreditation on quality enhancement (Andres et al., 2019; Schmaltz et al., 2011). Despotou et al., (2020), in a study among nurses in South Korea, found that accreditation has a positive impact on patient safety in tertiary care. Accreditation could bring continuity of quality patient care, and human resource management processes improved across time(Greenfield & Braithwaite, 2009). Sheikh (2017) found that accreditation has a positive impact on the satisfaction of the pharmacy department in a private tertiary care hospital at Secunderabad, Telangana State, India. Similarly, Camillo et al., (2016) found that accreditation is a favourable system for quality management in the public service because it promotes the development of professional skills and improves cost management, organisational structure, management of assistance and perception of job pride/satisfaction. Accreditation is an interface that strengthens trust between medical institutions and patients, especially in undeveloped countries (Spasojevic and Susic, 2011). Williams et al., (2017) compared the quality ratings of accredited and non-accredited nursing homes and found that accreditation is a significant predictor of quality enhancement.

Nonetheless, many studies established that accreditation was not associated with considerable improvement in healthcare delivery(Bogh et al., 2015; Lam et al., 2018; Rosenberg et al., 2016). Accreditation mainly emphasis on improving structural factors and clinical processes rather than improving patient outcomes. Al Otaibi, Kattan and Nabil, (2020) found the reverse effect of CBAHI (Saudi Central Board for Accreditation of Healthcare Institutions) on patient safety and failed to create total quality management.

Although there are many studies on accreditation structure, performance and patient satisfaction, the results are contradictory and inconclusive. A holistic comparative study of accreditation impact in public healthcare facilities using various quality dimensions is rare. Moreover, previous studies have almost focused exclusively on accredited hospitals with pre-test and post-test and measured the outcome without having a comparison group and vice versa. More importantly, there has been no previous evidence for studies comparing the effectiveness of national and state level accreditation programmes. In this context, this paper identifies nine dimensions of quality healthcare delivery and examines its impact on patient satisfaction (outcome) both in accredited (NABH and KASH) and non-accredited public healthcare settings of Kerala. This study results will throw light into the impact of implementing national and state level accreditation programmes. To this end, the following two hypotheses are formulated and tested using appropriate statistical techniques.

1. *Quality healthcare delivery enhances patient satisfaction in accredited hospitals in Kerala*.
2. *NABH accreditation creates more impact than KASH in various dimensions of quality healthcare delivery in Kerala*.

A conceptual framework is developed with ten latent constructs on the basis of the review of literature (see Figure 1).

**Fig.1.**
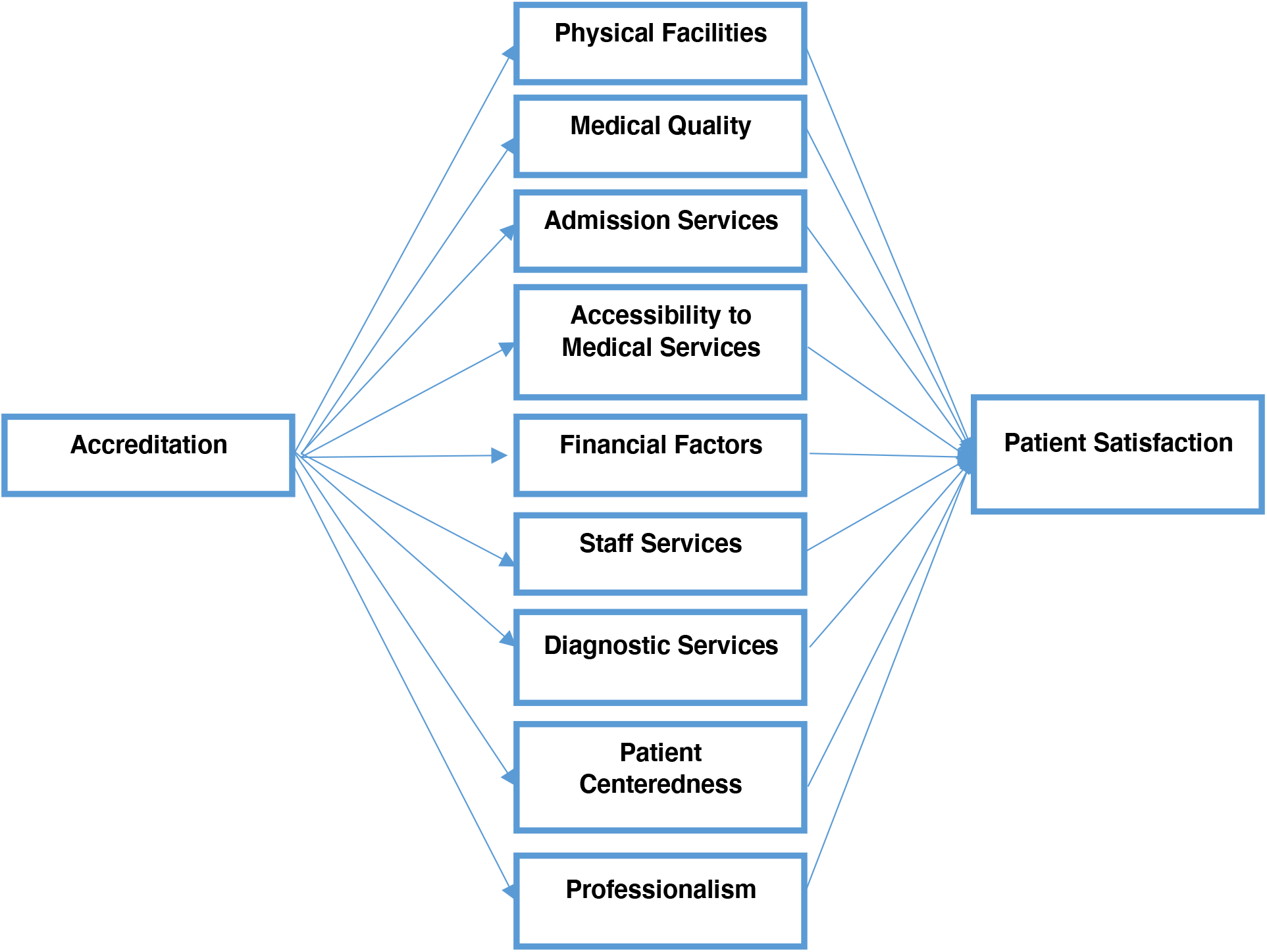
Conceptual Framework

## 2. METHODS

It is a cross sectional study drew on a positivist approach. Kerala healthcare model functions through a three-tier system, and therefore, the research used stratified random sampling where four strata – GHs, W&C hospitals, ‘Taluk Hospitals’ (THs)/THQHs and CHCs- are selected randomly from Southern, Central and Northern regions of Kerala. The study’s target population was in-patients admitted to medical wards at public hospitals, both accredited and non-accredited. Informed consent was obtained from them after describing the nature of the survey and prior permission to collect data from IP wards was obtained from the Department of Health Services (DHS) Kerala. In-patients aged 16 years or older and able to speak Malayalam or English language were included in the study. Being primary care facilities, PHCs were excluded from the study due to the lack of an acceptable number of in-patients.

**Table 1.**
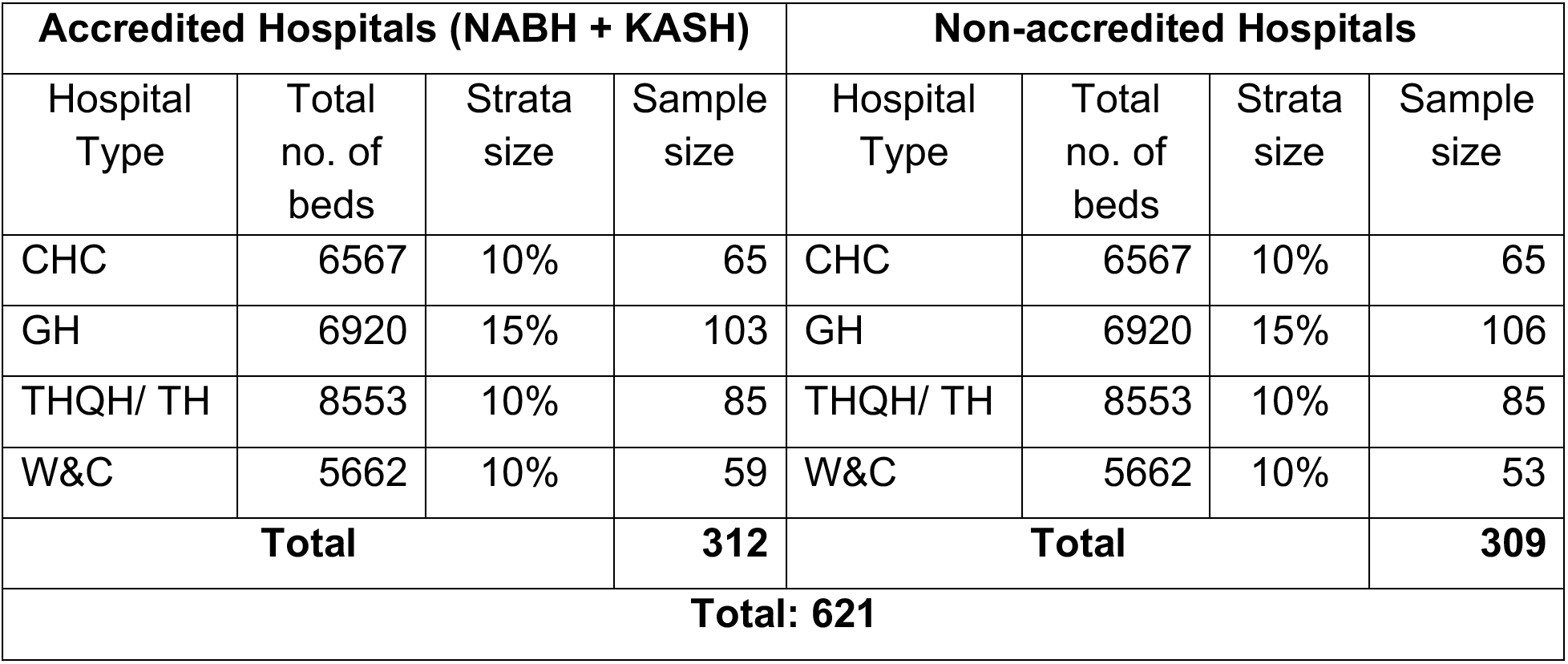
Sampling

| Accredited Hospitals (NABH + KASH) |  |  |  | Non-accredited Hospitals |  |  |  |
| --- | --- | --- | --- | --- | --- | --- | --- |
| Hospital Type | Total no. of beds | Strata size | Sample size | Hospital Type | Total no. of beds | Strata size | Sample size |
| CHC | 6567 | 10% | 65 | CHC | 6567 | 10% | 65 |
| GH | 6920 | 15% | 103 | GH | 6920 | 15% | 106 |
| THQH/ TH | 8553 | 10% | 85 | THQH/ TH | 8553 | 10% | 85 |
| W&C | 5662 | 10% | 59 | W&C | 5662 | 10% | 53 |
| Total |  |  | 312 | Total |  |  | 309 |
| Total: 621 |  |  |  |  |  |  |  |

To get a valid number of samples, 10% of the number of beds from each stratum was included in the study except GHs (15%) where there is only one GH accredited in Kerala (See Table 1). A total of 760 questionnaires were circulated in the In-Patient wards, of which 621 (82%) were valid for analysis (312 from accredited and 309 from non-accredited) which is considered sufficient to represent a large population Saunders, Lewis, and Thornhill, 2009).

This study was conducted from July 2017 to July 2018 using a questionnaire with 60 items covering ten constructs by adopting previous critical studies and models in the area(Amin and Nasharuddin, 2013; Darwazeh, 2011; Joseph, 2012; Kang et al., 2012; Lam, 1997; Legido-Quigley et al., 2008; Mosadeghrad, 2012; Pai and Chary, 2016; Peprah, 2014; Tashkandi, Hejazi, and Lingawi, 2017; Zineldin, 2006). The five dimensions of SERVQUAL model (Tangibles, Reliability, Responsiveness, Assurance and Empathy) and the Donabedian’s Structure-Process-Outcome (SPO) model are also imbibed in the chosen constructs. The first part of the questionnaire comprised of demographic information such as age, gender, educational level, marital status, employment status and the reason for hospital selection. The second section included statements to measure patients’ opinion on healthcare received by them on a 5-point scale (1=Strongly Agree to 5=Strongly Disagree)(Gursoy & Rutherford, 2004). The variables were grouped under ten constructs, namely Physical Facility (14 items), Admission Services (2 items), Patient centeredness (7 items), Accessibility of Medical Care (5 items), Financial Matters (5 items), Professionalism (4 items), Staff Services (4 items), Medical Quality (4 items), Diagnostic Services (2 items) and Patient Satisfaction (5 items). Initially the questionnaire was developed in the English language and then subsequently translated into Malayalam. Based on the result from the pilot test, slight changes were made in few questions. The validity of the questionnaire was evaluated based on content validity and expert opinion. The constructs wise Cronbach’s Alpha value was higher than the guideline value of 0.6. Simple statistical techniques like descriptive statistics, t-test, and Kruskal Wallis tests have been undertaken for data analysis.

## 3. RESULTS

### 3.1. Baseline socio-demographic variables

Patients seek healthcare in government hospitals are mainly females, koolies (daily workers) and students and majority belong to < Rs. 5000 income group. The striking point is that, in accredited hospitals, only 2.2% of the patients are bothered about the accreditation status while choosing the hospital. ‘Free treatment’ is the dominant push factor for them (see Table 2).

**Table. 2:**
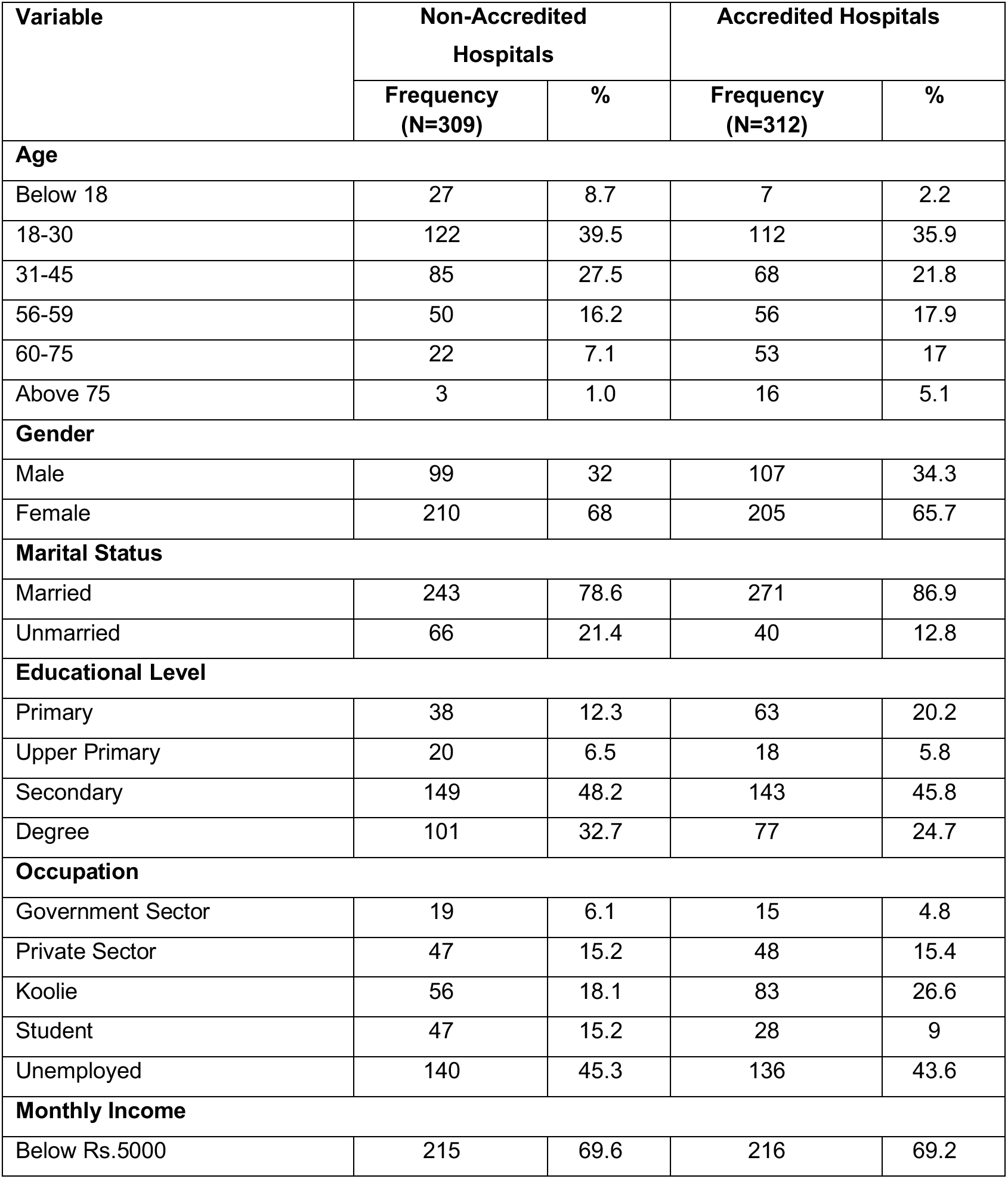
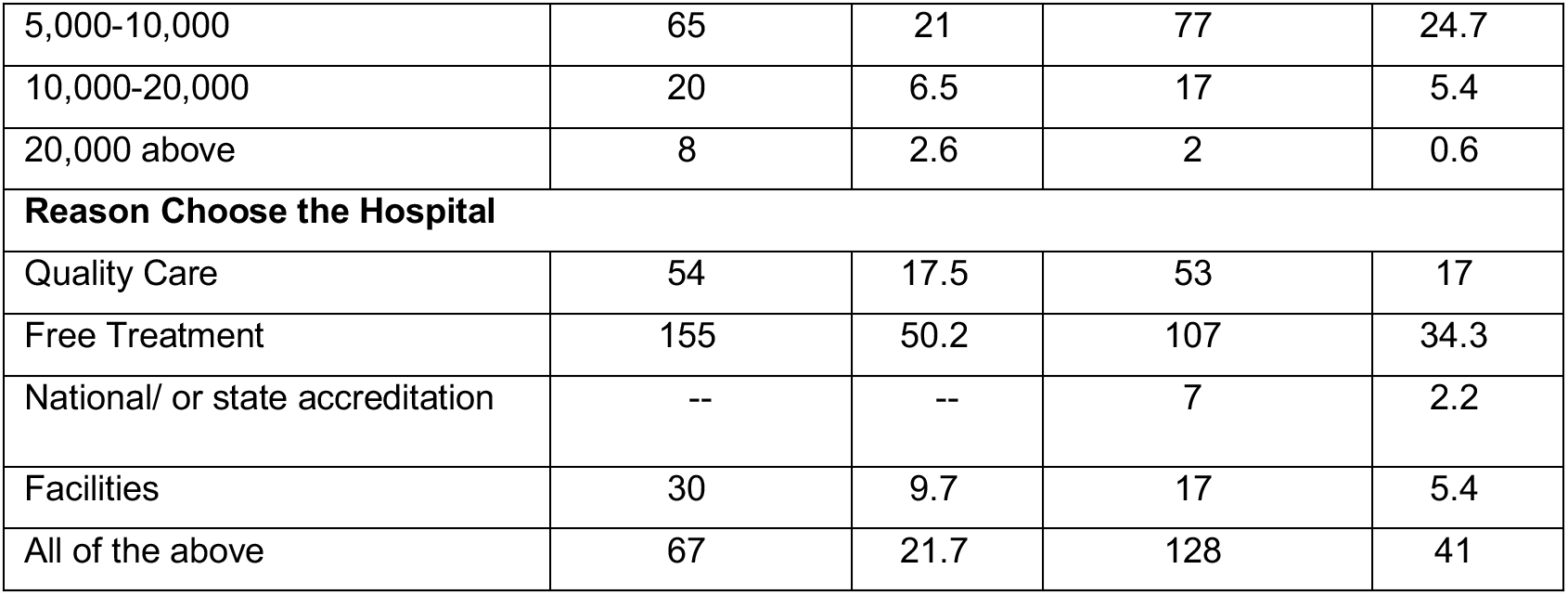
Baseline socio-demographic variables

### 3.2 Dimensions of Quality Healthcare Delivery

Patient Satisfaction (M=4.28) and Admission Services (M=4.27) get almost identical scores in both accredited and nonaccredited hospitals (see Table 3). The mean score of all the constructs of accredited hospitals is slightly higher than or equal to non-accredited except the ‘Medical Quality’ construct (Accredited M=4.44±0.9582; Non-accredited M=4.52±1.8519). However, the mean difference is statistically significant only for ‘Physical Facility’ (t (619) =-3.342, p=.001), Patient Centeredness (t (619) =-2.386, p=.017), Accessibility of Medical Services (t (619) = −2.968, p=.003) and Diagnostic Services (t (619) = −2.441, p=.015).

**Table 3.** Comparison of accredited vs non-accredited hospitals: t-test results for constructs of dimensions of quality healthcare delivery.

| Constructs | Dimensions of Quality Healthcare Delivery |  |  |  |  |  |
| --- | --- | --- | --- | --- | --- | --- |
|  | Status | Mean | SD | t- value | Df | p-value |
| Physical Facility | Non-accredited | 4.00 | 0.9729 | -3.342 | 619 | 0.001 |
|  | Accredited | 4.26 | 0.9785 |  |  |  |
| Admission Services | Non-accredited | 4.27 | 2.1237 | -0.025 | 607 | 0.980 |
|  | Accredited | 4.27 | 0.8780 |  |  |  |
| Patient Centeredness | Non-accredited | 4.25 | 0.7405 | -2.386 | 619 | 0.017 |
|  | Accredited | 4.41 | 0.9061 |  |  |  |
| Accessibility of Medical Services | Non-accredited | 3.68 | 0.9530 | -2.968 | 619 | 0.003 |
|  | Accredited | 3.93 | 1.1055 |  |  |  |
| Financial Factors | Non-accredited | 3.90 | 1.1313 | -0.595 | 619 | 0.451 |
|  | Accredited | 3.98 | 2.0157 |  |  |  |
| Professionalism | Non-accredited | 4.14 | 0.9954 | -1.003 | 619 | 0.552 |
|  | Accredited | 4.23 | 1.3168 |  |  |  |
| Staff Services | Non-accredited | 4.20 | 0.7447 | -1.075 | 618 | 0.316 |
|  | Accredited | 4.29 | 1.2648 |  |  |  |
| Medical Quality | Non-accredited | 4.52 | 1.1895 | 0.906 | 619 | 0.365 |
|  | Accredited | 4.44 | 0.9582 |  |  |  |
| Diagnostic Services | Non-accredited | 3.63 | 1.8519 | -2.441 | 619 | 0.015 |
|  | Accredited | 4.01 | 1.9916 |  |  |  |
| Patient Satisfaction | Non-accredited | 4.28 | 0.9478 | -0.051 | 619 | 0.959 |
|  | Accredited | 4.28 | 0.9195 |  |  |  |
| Accredited Hospital (N=312) |  | Non-accredited Hospital (N=309) |  |  |  |  |

The correlation analysis of the accredited hospitals indicated a significant positive association between patient satisfaction and the chosen constructs overarching various quality dimensions using Spearman’s correlation Coefficient (see Table.4). Here, the dependent variable patient satisfaction is correlated significantly with Physical Facilities (Spearman’s *ρ* = .702; p < 0.05), Admission Services (Spearman’s *ρ* = .506; p < 0.05), Patient Centeredness (Spearman’s *ρ* = .666; p < 0.05), Accessibility of Treatment (Spearman’s *ρ* = = .706; p < 0.05), Financial Matters (Spearman’s *ρ* = .531; p < 0.05), Professionalism (Spearman’s *ρ* = .704; p < 0.05), Staff Services (Spearman’s *ρ* = .747; p < 0.05), Medical Quality (Spearman’s *ρ* = .703; p < 0.05) and Diagnostic Services (Spearman’s *ρ* = .720; p < 0.05) and hence the above hypothesis is confirmed. However, the Kruskal-Wallis test indicated that there is no significant positive association between Accreditation and Patient Satisfaction (χ^2^=3.857, p<0.05).

**Table 4.** Hypothesis testing – dimensions of quality healthcare delivery enhances patient satisfaction in accredited hospitals

| Sl. No | Hypothesis(N=309) | Spearman's rho (r) | p-value |
| --- | --- | --- | --- |
| 1 | Physical Facilities- Patient satisfaction | 0.702 | 0.000 |
| 2 | Admission Services- Patient satisfaction | 0.506 | 0.000 |
| 3 | Patient Centeredness- Patient satisfaction | 0.666 | 0.000 |
| 4 | Accessibility of Treatment- Patient satisfaction | 0.706 | 0.000 |
| 5 | Financial Matters- Patient satisfaction | 0.531 | 0.000 |
| 6 | Professionalism- Patient satisfaction | 0.704 | 0.000 |
| 7 | Staff Services- Patient satisfaction | 0.747 | 0.000 |
| 8 | Medical Quality- Patient satisfaction | 0.703 | 0.000 |
| 9 | Diagnostic Services- Patient satisfaction | 0.720 | 0.000 |
| p<0.05; N=312 |  |  |  |

### 3.3 Type of accreditation (NABH vs. KASH)

For NABH hospitals, the mean score of only four constructs (Patient Centeredness, Accessibility of Medical Services, Professionalism and Staff Services) is higher (see Table 5). The mean score of five constructs (Physical Facility, Admission Services, Medical Quality, Financial Matters and Diagnostic Services) is higher in KASH accredited hospitals. Again, it shows that the differences were statistically not significant for six constructs (Patient Centeredness, Accessibility of medical Services, Professionalism, Staff Services, Medical Quality and Diagnostic Services). The mean score of the construct ‘Patient Satisfaction’ in NABH accredited hospital (M=4.27±0.67874) is lower than that of KASH accredited hospital (M=4.30±1.25417) even though the difference is statistically not significant (t (310) =-.257, p=.798).

**Table 5:** Comparison of differences of mean scores of constructs between NABH and KASH accredited hospitals

| Constructs | Type of the Hospital | Mean | SD | t value | Df | p-value |
| --- | --- | --- | --- | --- | --- | --- |
| Physical Facility | NABH | 4.15 | 0.59499 | -2.878 | 310 | 0.004 |
|  | KASH | 4.48 | 1.4224 |  |  |  |
| Admission Services | NABH | 4.16 | 0.87199 | -3.072 | 298 | 0.002 |
|  | KASH | 4.48 | 0.85507 |  |  |  |
| Patient Centeredness | NABH | 4.47 | 0.95419 | 1.415 | 310 | 0.158 |
|  | KASH | 4.31 | 0.80387 |  |  |  |
| Accessibility of medical Services | NABH | 3.94 | 1.0983 | 0.335 | 310 | 0.738 |
|  | KASH | 3.90 | 1.1235 |  |  |  |
| Financial Factors | NABH | 3.67 | 1.2780 | -3.708 | 310 | 0.000 |
|  | KASH | 4.54 | 2.8545 |  |  |  |
| Professionalism | NABH | 4.24 | 1.5398 | 0.253 | 310 | 0.800 |
|  | KASH | 4.20 | 0.74653 |  |  |  |
| Staff Services | NABH | 4.30 | 1.11094 | 0.273 | 310 | 0.785 |
|  | KASH | 4.26 | 1.51584 |  |  |  |
| Medical Quality | NABH | 4.43 | 1.12193 | -0.269 | 310 | 0.788 |
|  | KASH | 4.46 | 0.53802 |  |  |  |
| Diagnostic Services | NABH | 3.91 | 0.88508 | -1.219 | 310 | 0.224 |
|  | KASH | 4.20 | 3.14696 |  |  |  |
| Patient Satisfaction | NABH | 4.27 | 0.67874 | -0.257 | 310 | 0.798 |
|  | KASH | 4.30 | 1.25417 |  |  |  |

## 4. DISCUSSION

This study was conducted with the aim of examining the dimensions of quality healthcare delivery in accredited public healthcare institutions in Kerala. The analysis did not show any significant difference in patient satisfaction between accredited and non-accredited hospitals. Satisfaction is an expression of the patients’ overall judgment on the quality of care, including interpersonal aspects (Donabedian, 1980) and ‘how well’ the services provided to meet their needs and expectations (Haj-Ali et al., 2014). This study is validated by many earlier studies (Greenfield et al., 2008; Haj-Ali et al., 2014; Hayati et al., 2010; Heuer, 2004; Sack et al., 2010; Sack et al., 2011). However, the present study establishes that quality is marginally enhanced due to accreditation process.

Planned comparisons revealed that KASH accreditation has more impact on quality healthcare delivery and patient satisfaction which ties well with the previous study, wherein Lam et al., (2018) could not observe better patient experience at JCI accredited hospitals, and satisfaction was slightly worse compared with the level of satisfaction at state survey hospitals. The same study result was obtained by them over the consecutive years (2014 and 2015). This result is also congruent with the study of Greenfield & Braithwaite(2009) who found that organisations with different levels of accreditation, the performance showed varied rates of improvement. It is worth discussing the interesting fact that a contradictory result has been obtained in the case of healthcare dimension constructs of NABH and KASH, where NABH is a higher-order national level accreditation system. Patient satisfaction is lower in NABH than KASH, and that raises questions about the implementation of NABH programme, a long-drawn expensive process requiring many inspections over a period of three to five years.

Physical Facility and Diagnostic Services are significantly improved as the accreditation process largely focuses on infrastructure development (Haj-Ali et al., 2014; Sack et al., 2011). Accreditation eased doctor consultation and patient-doctor communication which is an important concern in hospital choice. However, accreditation could not bring remarkable changes in other areas requiring humane approach, therefore, not impacted in patient satisfaction. In addition, patients do not get any added financial advantage while seeking healthcare from an accredited public healthcare facility. This is not something to ignore when 42.25% of the population depends on public facility due to their financial insecurities, 69.4% belongs to <Rs.5000 income group and 66.85% are from the venerable section of the society (women, koolies (daily workers), and students) (Refer, Table.2). Maya (2016) noted that the costs of one hospitalisation episode (in-patient) in the private sector in Kerala is Rs.22, 989, which is 200 percent higher when compared to the costs in the public sector, Rs.11,065. The cost of an outpatient visit in the private sector is Rs.525, which was Rs.391 in the public sector. Obviously, it is crucial to strengthen the public sector when the overall healthcare costs in Kerala remain high.

The study results are steady with the earlier studies(Greenfield & Braithwaite, 2009; Sack et al., 2011; van Doorn – Klomberg et al., 2014) that hospital accreditation is not necessarily a contributor for quality care even though it has a reflection total quality enhancement. Further, these results go against the primary focus of the Twelfth Five Year Plan of Kerala (2012-17), which was quality care. However, the study shows that accreditation programs can facilitate continual and systematic improvement to hospitals sub-systems (Greenfield et al., 2019). It is important to recall the observation of WHO (2019, p.227) that despite the amount of money invested in the implementation of accreditation programs, evidence on cost-effectiveness is almost non-existent. To this end, Lam et al., (2018, p.227) suggested that “If we are to continue to use accreditation – and spend the substantial sums of money they require – then we should consider substantially rethinking our accreditation process.” In this context, all quality enhancement program must ensure the attainment of fundamental goals of health systems for health improvement and responsiveness to legitimate expectations of the patients (WHO, 2000) while ensuring cost-effectiveness. Viewing the successful Kerala healthcare model in preventing the epidemic outbreaks with its organised three tier public healthcare system, Kerala can upgrade its public health facilities qualitatively and quantitatively with an additional financial healthcare allocation and making use of Kerala’s dedicated and efficient healthcare workers.

The results demonstrate three major findings. First, accreditation is not an essential tool for bringing a remarkable improvement in quality healthcare delivery and, thereby, patient satisfaction. Second, accreditation can enhance the quality only through the sufficient allocation of funds, employing adequate and professional human resource(Yousefinezhadi et al., 2020), providing free treatment including diagnostic tests, adopting personalised approach through the effective implementation of the accreditation program. Third, Kerala healthcare institutions may favor KASH accreditation as an initial step towards quality enhancement than the national level NABH accreditation.

The data was collected at the same period of Nipah outbreak (2018 May)which caused geographical limitations and denial in accessing in-patients. To truly assess the impact of accreditation, the study requires more data from the same strata and, therefore, future research should further confirm these initial findings by including all accredited hospitals under study.

## 5. CONCLUSION

The evidence from the current study suggests that there is no significant impact of accreditation on patient satisfaction. Nevertheless, the different dimensions of healthcare are marginally improved through the accreditation process. As a step towards quality enhancement, accreditation programme should be monitored effectively to get the best results. Overall, these results suggest that it is vital to train and motivate all healthcare workers with an attitude of patient-centeredness, work efficiency, better recognition of patient’s needs with promptness, bridging the communication gap between the caregivers and patients for effective healthcare delivery.

## Data Availability

Data submission is suubjected to the funding agency approval.

## Author Statements

### Ethical approval

The entire study was conducted in accordance with the guidelines and ethical approval received from the Department of Health, Government of Kerala and the Indian Council for Social Science Research (ICSSR). Informed consent was obtained from the respondents before collecting their data. A prior written permission was also obtained from the Directorate of Health Services, Kerala to access in-patients in medical wards.

